# Investigating low birthweight and preterm birth as potential mediators in the relationship between prenatal infections and early child development: A linked administrative health data analysis

**DOI:** 10.1101/2024.03.13.24304219

**Authors:** Iain Hardie, Aja Murray, Josiah King, Hildigunnur Anna Hall, Kenneth Okelo, Emily Luedecke, Louise Marryat, Lucy Thompson, Helen Minnis, Michael Lombardo, Philip Wilson, Bonnie Auyeung

## Abstract

**Background:** Prenatal infections are associated with childhood developmental outcomes such as reduced cognitive abilities, emotional problems and other developmental vulnerabilities. However, there is currently a lack of research examining whether this arises due to potential intermediary variables like low birthweight or preterm birth, or due to some other mechanisms of maternal immune activation arising from prenatal infections.

**Methods:** Administrative data from the National Health Service (NHS) health board of Greater Glasgow & Clyde, Scotland, was used, linking birth records to hospital records and universal child health review records for 55,534 children born from 2011-2015, and their mothers. Causal mediation analysis was conducted to examine the extent to which low birthweight and preterm birth mediate the relationship between hospital-diagnosed prenatal infections and having developmental concern(s) identified by a health visitor during 6-8 week or 27-30 month child health reviews.

**Results:** Model estimates suggest that 5.18% [95% CI: 3.77-7.65%] of the positive association observed between hospital diagnosed prenatal infections and developmental concern(s) was mediated by low birthweight, whilst 7.37% [95% CI: 5.36-10.88%] was mediated by preterm birth.

**Conclusion:** Low birthweight and preterm birth appear to mediate the relationship between prenatal infections and childhood development, but only to a small extent. Maternal immune activation mechanisms unrelated to low birthweight and preterm birth remain the most likely explanation for associations observed between prenatal infections and child developmental outcomes, although other factors (e.g. genetic factors) may also be involved.

## Introduction

Maternal health during pregnancy plays an important role in shaping later childhood development, and there is now a growing body of evidence suggesting that prenatal maternal infections are associated with early childhood developmental outcomes. Previous research has found associations between maternal infections during pregnancy and reduced cognitive abilities [1, 2], emotional difficulties [3], health visitor developmental concerns [4] and a range of other developmental vulnerabilities [5] in children. There is also a large body of evidence linking prenatal maternal infections to childhood diagnoses of neurodevelopmental conditions such as autism [6-8] and attention-deficit/hyperactivity-disorder (ADHD) [9], as well as to adulthood mental health conditions including schizophrenia [10] and bipolar disorder [11]. It has been postulated that this arises as a result of the immune response of mothers to prenatal infections, known as maternal immune activation (MIA), which creates a cascade of events affecting fetal brain development. This has been shown in animal models [12-14], and human studies are broadly consistent with this [15]. Consequently, MIA tends to be cited as the most likely reason for associations observed between prenatal maternal infections and childhood developmental outcomes in existing research [1-3, 5].

It is important to note, however, that MIA is not the only possible explanation for associations between prenatal infections and childhood developmental outcomes. In particular, one alternative explanation is that both prenatal infections and childhood developmental issues may be caused by genetic factors. A lot of antenatal pyelonephritis (bacterial kidney infection), for example, is associated with congenital structural abnormalities in the renal tract, many of which are heritable, and there are well-known associations between minor dysmorphic features and developmental delays [16, 17].

Nevertheless, if associations between prenatal infections and childhood development are caused by MIA rather than genetic factors, then MIA may exert its impact, in part, through some potential intermediary factors or ‘mediator variables’ (i.e. variables which appear in the causal sequence between two other variables [18]) such as low birthweight or preterm birth. This is demonstrated in Figure 1. Specifically, low birthweight and preterm birth may potentially be important intermediary or mediator variables in the association between prenatal infections and childhood development as: (a) research suggests some prenatal infections are associated with low birthweight and preterm birth [19-22], and (b) research suggests preterm birth and low birthweight are in turn associated with poorer child developmental outcomes [23-25]. Despite this, existing studies have tended to either ignore low birthweight and preterm birth from their modelling, or have included them in models but only as control variables rather than mediators [1-3, 5, 6].

**Figure 1.**
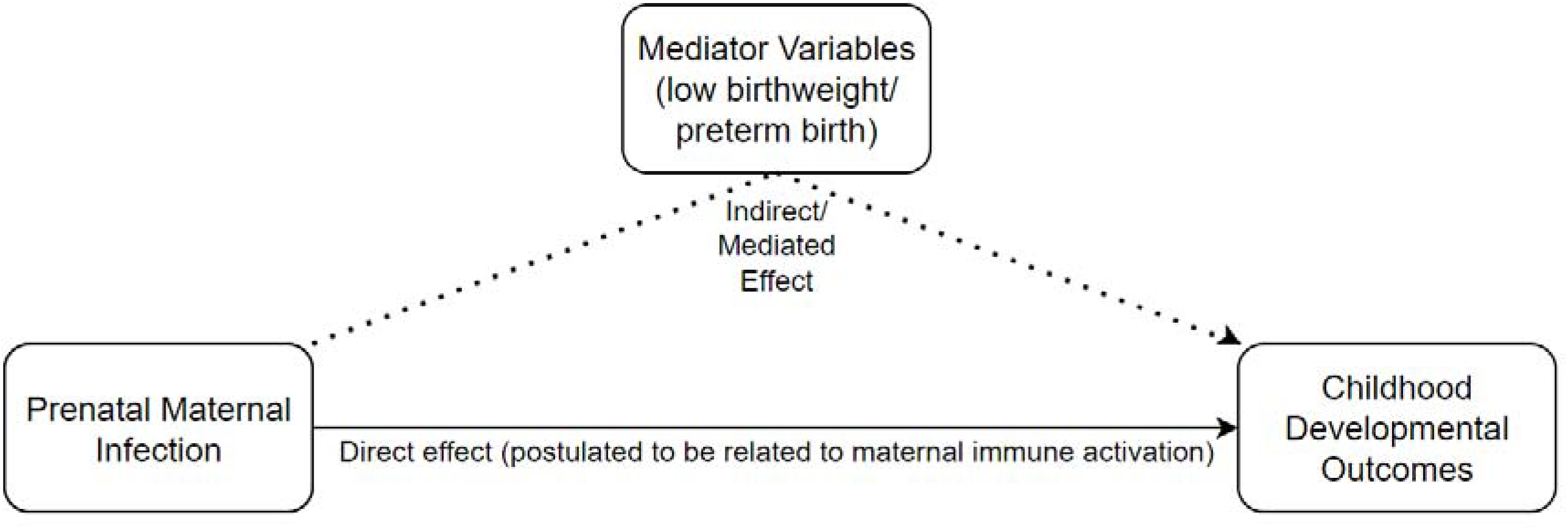
Diagram of the hypothesised relationship between prenatal infections, mediator variables and childhood developmental outcomes.

Whilst low birthweight and preterm birth are already considered factors that would flag risk of poorer developmental outcomes, better understanding of MIA mechanisms and how much of its impact is and is not mediated could help inform interventions in childhood development. In order to gain insight into this, the present study used a large linked administrative health dataset from the National Health Service (NHS) health board of Greater Glasgow & Clyde, Scotland, to examine the following research question:

> To what extent do low birthweight and preterm birth mediate the relationship between hospital-diagnosed prenatal maternal infections and childhood developmental outcomes, as measured during child health reviews at age 6-8 weeks and age 27-30 months?

## Methods

### Research design

A causal mediation analysis of a large linked administrative health dataset from NHS Greater Glasgow & Clyde, Scotland.

### Data and participants

The dataset was comprised of Scottish birth records, hospital records and routine child health review records. Participants were children born from 2011-2015 in NHS Greater Glasgow & Clyde, Scotland, and their mothers. Full details of the inclusion criteria, and number of exclusions, are provided in Figure 2. The final number of participants was 55,534 child-mother pairs.

**Figure 2.**
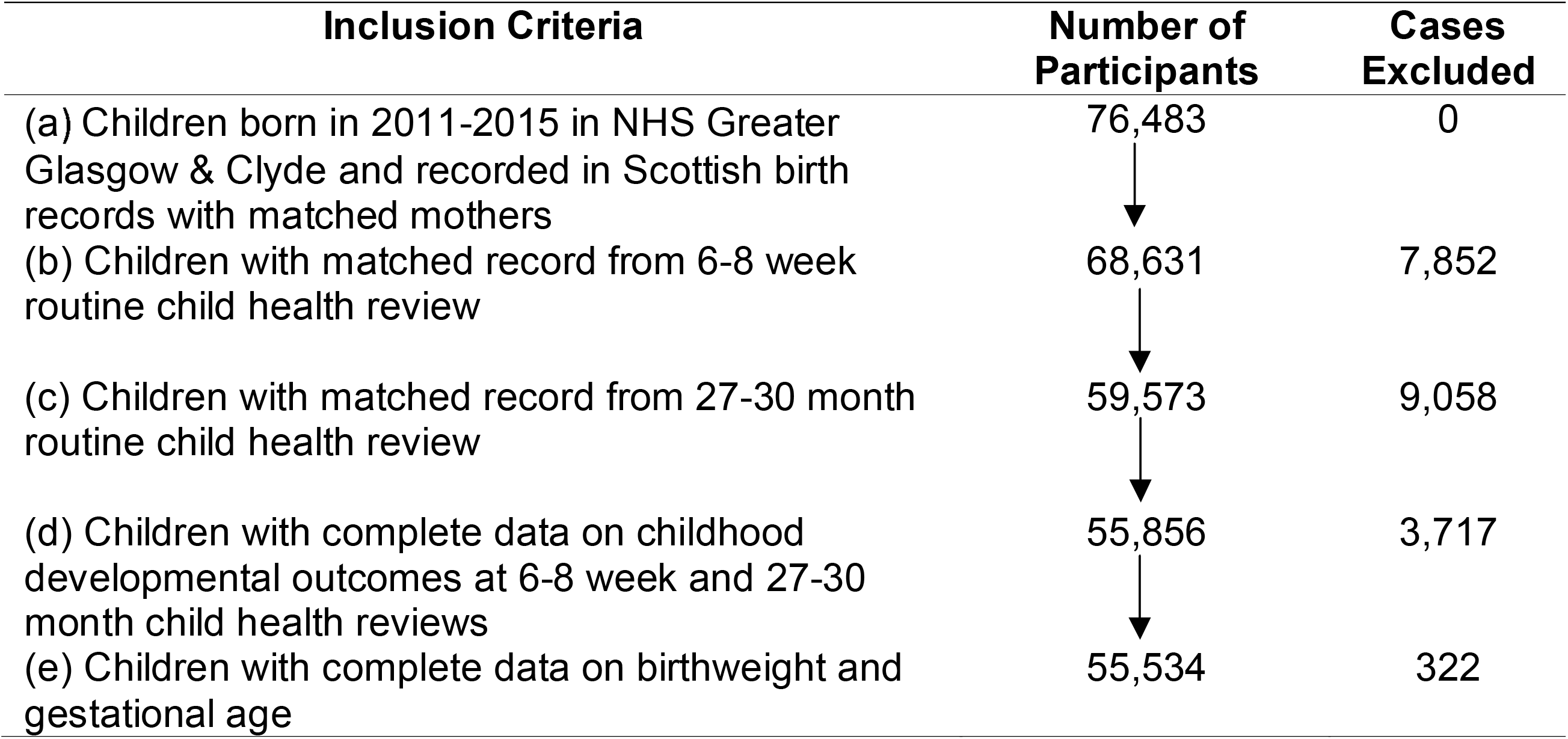
Flowchart of inclusion criteria, exclusions and number of participants.

### Measures

#### Outcome variables

In Scotland, all pre-school children are eligible to receive routine health reviews through the Scottish Government’s ‘Universal Health Visiting Pathway’. These are conducted in order to review children’s health, development, and wellbeing, and to provide advice and parenting support [26, 27].

Here, data from the 6-8 week and 27-30 month versions of these health reviews were used to create a binary outcome variable indicating whether any (i.e. at least one) childhood developmental concerns were identified by health visitors during one of these reviews (coded: 0=‘No’, 1=‘Yes’). This variable is referred to herein as ‘developmental concern(s)’, and could relate to any of the following developmental outcomes: gross-motor-skills development, hearing-communication development, and vision-social-awareness development (assessed at 6-8 week reviews), and personal-social development, emotional-behavioural-attention development, and speech-language-communication development (assessed at 27-30 month reviews). Developmental concerns were defined as cases where health visitors categorised a developmental outcome as either ‘a concern’, ‘abnormal’ or ‘doubtful’ (doubtful indicates a possible or likely concern/abnormality). These judgements were made by health visitors through: (a) eliciting parental concerns, (b) making structured observations of the child and/or (c) using validated developmental questionnaires [26].

Additional sensitivity analysis (outlined below) also included an additional outcome measure, ‘number of developmental concerns’. This was a numeric variable indicating the total number (from 0-6) of developmental concerns identified by health visitors.

#### Explanatory variable

The main explanatory variable was a binary measure of ‘hospital-diagnosed prenatal infections’ (coded: 0=‘No’, 1=‘Yes’). Here, hospital records from general or maternity hospital admissions [28, 29] were used to identify cases where mothers were diagnosed with at least one infection during their pregnancy. This was done using ‘International Classification of Diseases 10’ (ICD10) codes for infections. A full list of all the ICD10 codes included is provided in Table 1. A list of the top 20 most prevalent specific (i.e. four digit) ICD10 infection codes is also provided in supplemental appendix 1.

**Table 1.** List of ICD10 codes included in ‘hospital-diagnosed maternal infections during pregnancy’ variable.

| **ICD10 Codes** | **Type of Infections Included** |
| --- | --- |
| A/B | virus infections; bacterial infections; sexually transmitted infections/diseases |
| G0/G531/G630/ G940 | meningitis; cranial nerve infections; polyneuropathy infections; hydrocephalus infections |
| H0/H1/H20/H22/H320/H440/H441/H451/H481 | eye infections |
| I30/I310/I311/I32/I33/I38/I39/I40/I41/I430/I520/I521/I681/I980/I981 | pericarditis; endocarditis; myocarditis; heart infections; artery infections; cardiovascular infections |
| J0/J1/J2/J31/J32/J35/J36/J37/J39/J40/J41/J42/J440/J85/J86 | sinusitis; tonsillitis; laryngitis; common cold; influenza; pneumonia; respiratory infections; bronchitis; rhinitis; pharyngitis; peritonsillar abscess; pharynx infections; pulmonary disease with infection; gangrene/lung abscess; pyothorax |
| K04/K05/K102/K112/K113/K122/K140/K35/K61/K630/K65/K67/K75/K770/K81/K930 | periodontitis; pulpitis; gingivitis; necrosis; jaw inflammation/infection; salivary gland infection; cellulitis/mouth abscess; glossitis; appendicitis; anal infection/abscess; intestine abscess/infection; peritonitis; liver infection; cholecystitis; tuberculous of intestine |
| L0 | skin infections |
| M0 | septic arthritis |
| N080/N10/N11/N12/N22/N290/N291/N30/N330/N34/N390/N61/N7 | glomerular disorders involving infection; interstitial nephritis; urinary calculus infection; kidney infection; cystitis; urethritis; urinary tract infection unspecified; infection in breast; pelvic/vaginal/vulva infection or inflammation |
| O23/O98 | infections/infectious disease associated with pregnancy |
| R50 | fever |
| T880 | Infection following immunisation |

Additional sensitivity analysis (outlined below) also included an alternative measure of the ‘hospital-diagnosed prenatal infections’, in which infections occurring in the month of childbirth were excluded.

#### Mediator variables

‘Low birthweight’ and ‘preterm birth’ were used as mediator variables. Low birthweight was defined as a binary variable using the World Health Organisation (WHO) definition [30] of weighing <2,500 grams (coded: 0=‘Not low birthweight’, 1=‘low birthweight’. Similarly, preterm birth was a binary variable defined using the WHO definition [31] of being born before 37 weeks of gestational age (coded: 0=‘Not preterm’, 1=‘Preterm’). Data on both low birthweight and preterm birth were taken from Scottish birth records and maternity hospital records.

#### Control variables

Five confounders/covariates were included as control variables. These were: (a) ‘sex of child’, which was a binary variable from birth records (coded: 0=‘Male’, 1=‘Female’), (b) ‘area-based deprivation’, which was a categorical variable indicating which Scottish Index of Multiple Deprivation (SIMD) quintile the child’s home postcode was in (coded: 1=most deprived, 2=more deprived, 3=medium deprived, 4=less deprived, 5=least deprived), (c) ‘maternal age’, which was a continuous variable, from maternity records, indicating the age of the child’s mother at time of childbirth, (d) ‘maternal prenatal smoking’, which was a binary variable, also from maternity records, indicating whether the child’s mother smoked during their pregnancy (coded: 0=‘No’, 1=‘Yes’), and (e) ‘maternal history of mental health hospital admission’, which was a binary variable, from mental health hospital records, indicating whether the child’s mother had any record of historic mental health hospital admissions, i.e. including all historic records up to and including 12 months after the birth of their child (coded: 0=‘No’, 1=‘Yes’).

### Analysis

First, descriptive analysis was conducted in order to ascertain total frequencies of all variables included in the analysis, as well as frequencies across childhood development, prenatal infections, and mediator variables.

Next, the mediation analysis was conducted to formally investigate whether the relationship between prenatal infections and childhood developmental concern(s) was mediated by the low birthweight or preterm birth measures. Causal mediation analysis, as a method, plays an important role in helping to determine how and why particular treatment effects arise by helping researchers to identify intermediate variables that occur in the pathway between a given treatment and a given outcome [32]. Moreover, causal mediation analysis, in general, has multiple advantages over conventional mediation methods, such as allowing for the evaluation of necessary assumptions to ascertain the causal role of the mediator variable of interest [33]. The analysis for the present study was specifically carried out using a ‘potential outcomes’ framework via the Stata/MP 16 ‘*mediation*’ package, which estimates mediation effects using Monte Carlo simulations [32, 34, 35]. This works by simulating predicted values of a specified mediator or outcome variable and then calculating appropriate quantities of interest, i.e. average causal mediation, direct effects and total effects [34]. Here, models were specified as follows using the ‘*mediation*’ package’s ‘medeff’ command [34, 36, 37]:

- Outcome: ‘developmental concern(s)’
- Treatment: ‘hospital-diagnosed prenatal infections’
- Mediators: ‘low birthweight’ and ‘preterm birth’ (in two separate models)
- Control variables: ‘sex of child’, ‘area-based deprivation’, ‘maternal age’, ‘maternal prenatal smoking’ and ‘maternal history of mental health hospital admission’

As outcome and mediator variables were binary measures, logit models were used. The same set of control variables, i.e. all those listed above, were used to estimate predicted values for both the mediators and outcome. The only exception was that the maternal history of mental health hospital admissions control variable was omitted from the mediator model because it includes admissions from the first year of the child’s life and so temporally could have occurred after low birthweight/preterm birth had already occurred. Specifying the models like this gave estimates of the total effect of hospital-diagnosed prenatal infections on developmental concern(s), as well as the: (a) average direct effect (ADE), (b) average causal mediated effect (ACME) for each mediator variable, and (c) percentage of the total effect that is mediated by each mediator variable.

In addition to the main analysis, two sensitivity analyses were conducted. Firstly, the main analysis was repeated using the ‘number of developmental concern(s)’ outcome variable, as this potentially provides more detail into the extent of developmental concerns than the binary measure. As number of concern(s) is numeric, OLS modelling was used here for the outcome model rather than logit modelling. Secondly, the main analysis was repeated using the alternative measure of hospital-diagnosed prenatal infections which excluded infections from the month of childbirth. This was carried out because infections from the month of childbirth included, for example, Group B Strep carrier cases and herpes simplex virus encephalitis picked up when mothers are in hospital for delivery. These may be harmful for babies but are unlikely to cause MIA. Therefore, removing them may provide better insight potential links between MIA and child development.

All analysis was conducted within Scotland’s National Safe Haven. Reporting is consistent with ‘REporting of studies Conducted using Observational Routinely collected health Data’ (RECORD) guidelines [38]. The project received ethical approval from the University of Edinburgh’s Philosophy, Psychology and Language Sciences (PPLS) research ethics committee (reference: 190-1718/2), and information governance approval from the NHS Scotland Public Benefit and Privacy Panel for Health and Social Care (HSC-PBPP) (reference: 1617-0314).

## Results

### Descriptive statistics

Participant descriptive statistics are provided in Table 2. Overall, 21.2% of children had developmental concern(s), and these were disproportionately prevalent among those exposed to hospital-diagnosed prenatal infection(s), those with low birthweight/born preterm, and also among those who were male, from deprived areas, and whose mother smoked during pregnancy or had a history of mental health hospital admissions. Meanwhile, 5.1% of mothers had records of hospital-diagnosed infection(s) during pregnancy and these were more prevalent among those whose child had low birthweight or was born preterm, as well as among those from deprived areas, who smoked during pregnancy or had a history of mental health hospital admissions. With regards to mediator variables, 7.0% of children in the dataset had low birthweight and 7.5% were born preterm. Table 2 also highlights that both preterm birth and low birthweight appeared to be more prevalent among those whose mothers’ smoked during pregnancy or had a history of mental health hospital admissions.

**Table 2.** Descriptive statistics of participants by childhood development, prenatal infections and mediator variables.

|  |  | **Childhood development** |  | **Prenatal infection(s)** |  | **Mediator variables** |  |  |  |
| --- | --- | --- | --- | --- | --- | --- | --- | --- | --- |
|  |  | One or more childhood developmental concerns identified by health visitor |  | Hospital-diagnosed prenatal infection(s) |  | Low birthweight |  | Preterm birth |  |
|  | *Total* | *No* | *Yes* | *No* | *Yes* | *No* | *Yes* | *No* | *Yes* |
| **Childhood development** |  |  |  |  |  |  |  |  |  |
| One or more childhood developmental concerns identified by health visitor |  |  |  |  |  |  |  |  |  |
| No | 43,771<br>(78.8%) |  |  | 41,703<br>(79.1%) | 2,842<br>(72.8%) | 41,022<br>(79.4%) | 2,749<br>(70.7%) | 40,809<br>(79.4%) | 2,962<br>(71.4%) |
| Yes | 11,763<br>(21.2%) |  |  | 10,989<br>(20.9%) | 774<br>(27.2%) | 10,625<br>(20.6%) | 1,138<br>(29.3%) | 10,574<br>(20.6%) | 1,189<br>(28.6%) |
| **Prenatal infections** |  |  |  |  |  |  |  |  |  |
| Hospital-diagnosed prenatal infection(s) |  |  |  |  |  |  |  |  |  |
| No | 52,692<br>(94.9%) | 41,703<br>(95.3%) | 10,989<br>(93.4%) |  |  | 49,102<br>(95.1%) | 3,590<br>(92.4%) | 48,887<br>(95.1%) | 3,805<br>(91.7%) |
| Yes | 2,842<br>(5.1%) | 2,068<br>(4.7%) | 774<br>(6.6%) |  |  | 2,545<br>(4.9%) | 297<br>(7.6%) | 2,496<br>(4.9%) | 346<br>(8.3%) |
| **Mediator variables** |  |  |  |  |  |  |  |  |  |
| Low birthweight |  |  |  |  |  |  |  |  |  |
| No | 51,647<br>(92.0%) | 40,809<br>(93.2%) | 10,625<br>(90.3%) | 49,102<br>(93.2%) | 2,545<br>(89.6%) |  |  | 49,945<br>(97.2%) | 1,702<br>(41.0%) |
| Yes | 3,887<br>(7.0%) | 2,962<br>(6.8%) | 1,138<br>(9.7%) | 3,590<br>(6.8%) | 297<br>(10.4%) |  |  | 1,438<br>(2.8%) | 2,449<br>(59.0%) |
| Preterm birth |  |  |  |  |  |  |  |  |  |
| No | 51,383<br>(92.5%) | 40,809<br>(93.2%) | 10,574<br>(89.9%) | 48,887<br>(92.8%) | 2,496<br>(87.8%) | 49,945<br>(96.7%) | 1,438<br>(37.0%) |  |  |
| Yes | 4,151<br>(7.5%) | 2,962<br>(6.8%) | 1,189<br>(10.1%) | 3,805<br>(7.2%) | 346<br>(12.2%) | 1,702<br>(3.3%) | 2,449<br>(63.0%) |  |  |
| **Confounder/covariates** |  |  |  |  |  |  |  |  |  |
| Sex of child |  |  |  |  |  |  |  |  |  |
| Male | 28,186 | 20,518 | 7,668 | 26,722 | 1,464 | 26,392 | 1,794 | 25,972 | 2,214 |
|  | (50.7%) | (46.9%) | (65.2%) | (50.7%) | (51.5%) | (51.1%) | (46.2%) | (50.5%) | (53.3%) |
| *Female* | 27,348<br>(49.3%) | 23,253<br>(53.1%) | 4,095<br>(34.8%) | 25,970<br>(49.3%) | 1,378<br>(48.5%) | 25,255<br>(48.9%) | 2,093<br>(53.8%) | 25,411<br>(49.5%) | 1,937<br>(46.7%) |
| Area-based deprivation |  |  |  |  |  |  |  |  |  |
| 1 (*most deprived*) | 22,117<br>(39.8%) | 16,353<br>(37.4%) | 5,764<br>(49.0%) | 20,787<br>(39.4%) | 1,330<br>(46.8%) | 20,469<br>(39.6%) | 1,648<br>(42.4%) | 20,489<br>(39.9%) | 1,628<br>(39.2%) |
| 2 (*more deprived*) | 10,261<br>(18.5%) | 7,981<br>(18.2%) | 2,280<br>(19.4%) | 9,713<br>(18.4%) | 548<br>(19.3%) | 9,492<br>(18.4%) | 769<br>(19.8%) | 9,434<br>(18.4%) | 827<br>(19.9%) |
| 3 (*medium deprived*) | 8,405<br>(15.1%) | 6,804<br>(15.5%) | 1,601<br>(13.6%) | 8,006<br>(15.2%) | 399<br>(14.0%) | 7,834<br>(15.2%) | 571<br>(14.7%) | 7,751<br>(15.1%) | 654<br>(15.8%) |
| 4 (*less deprived*) | 7,126<br>(12.8%) | 5,988<br>(13.7%) | 1,138<br>(9.7%) | 6,785<br>(12.9%) | 341<br>(12.0%) | 6,709<br>(13.0%) | 417<br>(10.7%) | 6,642<br>(12.9%) | 484<br>(11.6%) |
| 5 (*least deprived*) | 7,625<br>(13.7%) | 6,645<br>(15.2%) | 980<br>(8.3%) | 7,401<br>(14.5%) | 224<br>(7.9%) | 7,143<br>(13.8%) | 482<br>(12.4%) | 7,067<br>(13.8%) | 558<br>(13.4%) |
| Maternal age |  |  |  |  |  |  |  |  |  |
| Mean age | 29.6 years | 29.9 years | 28.5 years | 29.7 years | 28.4 years | 29.6 years | 29.9 years | 29.5 years | 30.1 years |
| Maternal prenatal smoking |  |  |  |  |  |  |  |  |  |
| No | 47,647<br>(85.5%) | 38,393<br>(87.7%) | 9,254<br>(78.7%) | 45,376<br>(86.1%) | 2,271<br>(79.9%) | 44,767<br>(86.7%) | 2,880<br>(74.1%) | 44,257<br>(86.1%) | 3,390<br>(81.7%) |
| Yes | 7,887<br>(14.2%) | 5,378<br>(12.3%) | 2,509<br>(21.3%) | 7,316<br>(13.9%) | 571<br>(20.1%) | 6,880<br>(13.3%) | 1,007<br>(25.9%) | 7,126<br>(13.9%) | 761<br>(18.3%) |
| Maternal history of mental health hospital admissions |  |  |  |  |  |  |  |  |  |
| No | 54,652<br>(98.4%) | 43,173<br>(98.6%) | 11,479<br>(97.6%) | 51,888<br>(98.5%) | 2,764<br>(97.3%) | 50,879<br>(98.5%) | 3,773<br>(97.1%) | 50,609<br>(98.5%) | 4,043<br>(97.4%) |
| Yes | 882<br>(1.6%) | 598<br>(1.37%) | 284<br>(2.4%) | 804<br>(1.5%) | 78<br>(2.7%) | 768<br>(1.5%) | 114<br>(2.9%) | 774<br>(1.5%) | 108<br>(2.6%) |

\*Notes: Table shows frequencies and percentages for categorical or binary variables, and mean values for continuous variable.

### Mediation analysis

Estimates from the mediation analysis, investigating whether low birthweight and preterm birth mediate the relationship between hospital-diagnosed prenatal infection(s) and developmental concern(s), are provided in Table 3. The results highlight that there do appear to be mediating effects for both low birthweight and preterm birth. In both cases, these apparent mediated effects were, however, fairly small, and appear to mediate only a small proportion of the total effect of hospital-diagnosed prenatal infections on developmental concern(s). Specifically, the results highlight a positive association between hospital-diagnosed prenatal infections and developmental concern(s) (β=0.046 [95% confidence interval (CI): 0.031-0.063] for total effect estimate). However, estimated mediation through low birthweight accounted for only 5.18% [95% CI: 3.77%-7.65%] of this total effect (with an ACME estimate of β=0.002 [95% CI: 0.001-0.004] compared to an ADE estimate of β=0.043 [95% CI: 0.029-0.060]), whilst estimated mediation through preterm birth accounted for just an estimated 7.37% [95% CI: 5.36%-10.88%] of it (with an ACME estimate of β=0.003 [95% CI: 0.002-0.005] compared to an ADE estimate of β=0.042 [95% CI: 0.028-0.059]).

**Table 3.** Estimates of causal mediation in the relationship between hospital-diagnosed prenatal infection(s) and having one or more developmental concern(s) identified at age 6-8 week or age 27-30 month child health reviews.

| **Mediator Variable** | **Total Effect ($\beta$)** | **Average Direct Effect ($\beta$)** | **Average Causal Mediated Effect ($\beta$)** | **% of Total Effect Mediated** |
| --- | --- | --- | --- | --- |
| Low Birthweight | 0.046<br>[0.031-0.063] | 0.043<br>[0.029-0.060] | 0.002<br>[0.001-0.003] | 5.18%<br>[3.77%-7.65%] |
| Preterm Birth | 0.046<br>[0.031-0.063] | 0.042<br>[0.028-0.059] | 0.003<br>[0.002-0.004] | 7.37%<br>[5.36%-10.88%] |

\*Notes: $\beta$ refers to beta coefficient. 95% confidence intervals are shown in square brackets underneath estimates. Models adjust for sex of child, area-based deprivation, maternal age, maternal prenatal smoking and maternal history of mental health hospital admission(s).

### Sensitivity analysis

Results of the sensitivity analysis, whereby the main analysis was repeated using the ‘number of developmental concern(s) variable as an additional outcome, are provided in supplemental appendix 2. The results are broadly consistent with the results of the main analysis.

Results of the sensitivity analysis in which the main analysis was repeated, but excluding hospital-diagnosed infections from the month of childbirth, are provided in supplemental appendix 3. Here, the estimated total effect of hospital-diagnosed prenatal infections on developmental concern(s) was higher, possibly reflecting the fact that MIA was more likely to have occurred when using this measure (given that many infections from during the month of childbirth would have been infections like Group B Strep carrier cases and herpes simplex virus encephalitis picked up when mothers are in hospital for delivery so would not have involved any MIA that could logically impact fetal brain development). Estimated mediation effects were broadly similar to the main analysis.

## Discussion

This causal mediation analysis study used linked administrative health data for NHS Greater Glasgow & Clyde, Scotland, to investigate whether low birthweight and preterm birth mediate the relationship between hospital-diagnosed prenatal infections and early childhood developmental concern(s). The results highlight that both low birthweight and preterm birth do appear to have mediating effects, but only to a small extent, mediating an estimated 5.18% [95% CI: 3.77-7.65%] and 7.37% [95% CI: 5.36-10.88%], respectively, of the relationship between hospital-diagnosed prenatal infections and developmental concern(s). Prior to the current analysis, several previous studies had linked prenatal infections to childhood developmental outcomes, including reduced cognitive abilities, socioemotional difficulties and developmental concerns and vulnerabilities [1-5], and posited that this most likely occurred due to the MIA arising from prenatal infections creating a cascade of events that impacted fetal brain development. The present findings support this view, whilst also adding to previous studies by suggesting that only a small proportion of the association between prenatal infections and child development may be related to the mediating role of low birthweight and preterm birth. Therefore, the majority of the association between prenatal infections and child development appears to occur due to other mechanisms, which are mostly likely mechanisms related to MIA. This study makes a unique contribution to knowledge in that it is the first known piece of research examining potential mediating factors in the relationship between prenatal infection and childhood developmental outcomes. A key strength of the analysis is that it was able to make use of a large linked administrative health dataset.

However, there are some limitations. Firstly, although the analysis controlled for measured confounders and uses a causal mediation framework, specifying the temporal sequencing of relationships, it is still difficult to disentangle the role of mediators from unmeasured confounding e.g. by genetic factors. Similarly, it is possible that the mechanisms causing MIA to be linked to low birthweight/preterm birth are the same mechanisms involved in causing poorer developmental outcomes. In particular, it is possible that MIA is associated with neurobiology (e.g. in a way which alters gene expression) which has two effects – one more proximal (inducing low birthweight/preterm birth) and one more distant (poorer developmental outcomes). If this is the case then mediation might make it look like MIA is having an effect on the more proximal outcome, and then the second indirect effect on developmental outcomes when in reality both could be linked to the similar prenatal effect but at different times. Another limitation is that this study’s analysis tested mediators independently of each other. Given that low birthweight and preterm birth are correlated with each other, this makes it difficult to disentangle their effects.

Finally, there are also some general limitations to using administrative health datasets like the one used in the present study’s analysis. For example, there can be nuances around how ICD10 codes are applied and interpreted by medical professionals or reviewers entering the codes. The present study used an extensive list of ICD10 infection codes (see Table 1), but cannot rule out that infections data may be affected by nuances in how ICD10 codes are applied e.g. variations over time or between how different medical professionals/reviewers use them. Moreover, administrative records exclude people who do not interact with health services [39-41]. People from deprived areas are known to be less likely to be included in the child health review data used for the present analysis [42], and as the present study was a complete case analysis this potentially makes the dataset less representative. Also, the analysis was limited to data on infections from hospitals only, so did not include any infections which were less severe or community treated.

To conclude, this causal mediation analysis study suggests that low birthweight and preterm birth may mediate the relationship between prenatal infections and childhood development, but only to a small extent. It remains the case that aspects of MIA unrelated to low birthweight and preterm birth are the most likely explanation for associations observed between prenatal infections and childhood developmental outcomes, although other factors, such as genetic factors, may be involved.

## Data Availability

The administrative health datasets used for this study are not publicly available but can be accessed via successfully applying to the NHS Scotland Public Benefit and Privacy Panel for Health and Social Care.

## Acknowledgements

Iain Hardie and Bonnie Auyeung were supported by the Economic and Social Research Council (ES/W001519/1) during the course of this work. Bonnie Auyeung was additionally supported by the European Union’s Horizon 2020 research and innovation programme under the Marie Skłodowska-Curie grant agreement No.813546, the Baily Thomas Charitable Fund TRUST/VC/AC/SG/469207686, the Data Driven Innovation. In addition, the authors would like to acknowledge the electronic Data Research and Innovation Service (eDRIS) team at Public Health Scotland for their support in obtaining approvals, the provisioning and linking of data and facilitating access to the National Safe Haven.

---

**Supplemental Appendix 1.** List of top 20 most commonly recorded maternal infections during pregnancy.

| **ICD10 Code** | **Full description of infection** | **Number of infections recorded** |
| --- | --- | --- |
| O234 | Unspecified infection of urinary tract in pregnancy | 463 |
| B951 | Strep group B, as the cause of diseases classified in other chapters | 274 |
| O239 | Other and unspecified genitourinary tract infection in pregnancy | 231 |
| O235 | Infections of genital tract in pregnancy | 218 |
| J22X | Unspecified acute lower respiratory infection | 178 |
| A491 | Streptococcal and enterococcal infection, site unspecified | 173 |
| N390 | Urinary tract infection, site unspecified | 166 |
| N771 | Vaginitis, vulvitis & vulvovaginitis in infectious and parasitic diseases | 88 |
| B373 | Candidiasis of vulva and vagina | 85 |
| O230 | Infections of kidney in pregnancy | 74 |
| B962 | E. Coli as the cause of diseases classified in other chapters | 61 |
| B349 | Viral infection, unspecified | 44 |
| B379 | Candiasis, unspecified | 43 |
| A099 | Gastroenteritis and colitis of unspecified origin | 39 |
| R509 | Fever, unspecified | 36 |
| B24X | Unspecified human immunodeficiency virus [HIV] disease | 32 |
| L022 | Cutaneous abscess, furuncle and carbuncle of trunk | 29 |
| N12X | Tubulo-interstitial nephritis, not specified as acute or chronic | 27 |
| B956 | Staph aureus as the cause of diseases classified to other chapters | 24 |
| B169 | Acute hepatitis B without delta-age | 23 |

Notes: these are the top 20 most commonly recorded ICD10 four digit codes for infections during pregnancy for mothers in the sample. ICD10 codes listed in both general hospital admission records and maternity hospital admission records are included. Each hospital admission record could include up to six ICD10 codes, i.e. one for main condition recorded and up to five for other conditions recorded. Main conditions and all other conditions that correspond to infections during pregnancy period count towards overall number of infections listed.

**Supplemental Appendix 2.** Sensitivity analysis estimates of causal mediation in the relationship between hospital-diagnosed prenatal infection(s) and number of developmental concern(s) identified at age 6-8 weeks or age 27-30 months child health reviews.

| **Mediator Variable** | **Total Effect ($\beta$)** | **Average Direct Effect ($\beta$)** | **Average Causal Mediated Effect ($\beta$)** | **% of Total Effect Mediated** |
| --- | --- | --- | --- | --- |
| Low Birthweight | 0.101<br>[0.075-0.129] | 0.096<br>[0.071-0.123] | 0.005<br>[0.003-0.007] | 4.59%<br>[3.59%-6.15%] |
| Preterm Birth | 0.101<br>[0.075-0.129] | 0.094<br>[0.069-0.121] | 0.007<br>[0.005-0.009] | 6.77%<br>[5.30%-9.09%] |

\*Notes: $\beta$ refers to beta coefficient. 95% confidence intervals are shown in square brackets underneath estimates. Models adjust for sex of child, area-based deprivation, maternal age, maternal prenatal smoking and maternal history of mental health hospital admission(s).

**Supplemental Appendix 3.** Sensitivity analysis estimates of causal mediation in the relationship between hospital-diagnosed prenatal infection(s) and having one or more developmental concern(s) identified at age 6-8 week or age 27-30 month child health reviews (infections in month of childbirth excluded)

| **Mediator Variable** | **Total Effect ($\beta$)** | **Average Direct Effect ($\beta$)** | **Average Causal Mediated Effect ($\beta$)** | **% of Total Effect Mediated** |
| --- | --- | --- | --- | --- |
| Low Birthweight | 0.057<br>[0.037-0.081] | 0.055<br>[0.035-0.078] | 0.003<br>[0.001-0.004] | 5.48%<br>[3.48%-7.76%] |
| Preterm Birth | 0.057<br>[0.037-0.081] | 0.054<br>[0.034-0.077] | 0.003<br>[0.002-0.005] | 5.85%<br>[4.16%-9.01%] |

\*Notes: $\beta$ refers to beta coefficient. 95% confidence intervals are shown in square brackets underneath estimates. Models adjust for sex of child, area-based deprivation, maternal age, maternal prenatal smoking and maternal history of mental health hospital admission(s).

